# Vascular differences between glioblastoma *IDH-wildtype* and astrocytoma *IDH*-mutant grade 4 at imaging and transcriptomic level

**DOI:** 10.1101/2022.06.20.22276639

**Authors:** María del Mar Álvarez-Torres, Adolfo López-Cerdán, Maria de la Iglesia Vayá, Elies Fuster-Garcia, Francisco García-García, Juan M García-Gómez

## Abstract

A global agreement in Central Nervous System (CNS) tumors classification is essential in order to decide treatment correctly, predict prognosis, evaluate treatment response, compare outcomes, and select adequate patients for clinical trials at international level.

The last update of the World Health Organization (WHO) of CNS tumor classification and grading 2021 considered for the first time glioblastoma *IDH*-wildtype and astrocytoma *IDH*-mutant grade 4 as different tumors. *IDH* mutation produces a metabolic reprogramming of tumor cells, thus affecting the processes of hypoxia and vascularity. The differences in the aggressiveness of these gliomas, which affect patient survival, are evident, with a clear advantage for those patients who present *IDH* mutated tumors.

Despite the clinical relevance of *IDH* mutation, current protocols do not include full sequencing for every patient. Alternative biomarkers could be useful and complementary to get a more reliable classification. In this sense, MRI-perfusion biomarkers, such as relative cerebral blood and flow (rCBV and rCBF), could be key because they are non-invasive, can be used from the presurgical moment, and do not suppose additional economical or effort costs.

The main purpose of this work is to evaluate and validate the association between MRI-DSC biomarkers and *IDH* mutation status in high-grade astrocytomas. In addition, to get a deeper understanding of the vascular differences, we aim to study the transcriptomic bases of these vascular differences between these two high-grade astrocytomas.

## Introduction

Astrocytomas grade 4 includes two different tumors since 2016, when the *CNS tumors classification and grading of the WHO* incorporated new entities according to both histology and molecular features^1,2^. This difference is maintained in the new WHO of CNS 2021^1^, and in addition, it is accentuated, having also changed its nomenclature to glioblastoma *IDH*-wildtype and astrocytoma *IDH*-mutant grade 4 (instead of only *glioblastoma*). *G*lioblastoma *IDH*-wildtype represents about 95 % of astrocytomas grade 4 and predominates in patients over 55 years of age. Astrocytoma *IDH*-mutant grade 4 (about 5 % of cases) is more frequent in younger patients or in those with a history of prior lower grade diffuse glioma^1–3^.

Relevant differences are found between these two astrocytoma types at the clinical level, since they cause different patient survival rates^3–6^ as well as distinct therapy responses^7–9^. Therefore, it is evident that an early-stage classification considering the *IDH* mutation is necessary to get an adequate prognostic evaluation and a more personalized treatment of patients with astrocytomas grade 4.

Despite its important role, the definition of full *IDH* evaluation can differ according to patient age^2^, clinical protocols, and centers. The absence of R132H *IDH1* and *IDH2* mutations in astrocytomas from patients over about 55 years old suggests that sequencing may not be needed in the setting of negative R132H *IDH1* immunohistochemistry in those patients. Since protocols do not include full sequencing for every patient, alternative biomarkers could be useful and complementary to get a more reliable classification. In this sense, MRI-based methodologies could be key because they are non-invasive, could be used from the presurgical moment, and do not suppose additional economical or effort costs.

In 2020, Hao Wu *et al*.^10^ evaluated the potential clinical impact of the Hemodynamic Tissue Signature (HTS) method^11^ for predicting *IDH* mutation status in patients with glioma tumors. They analyzed the association between the relative cerebral blood volume (rCBV) at the high angiogenic tumor (HAT) habitat and the *IDH* mutation status. A significantly decreased rCBV for the *IDH*-mutant group was found. They concluded that “the HTS method was proven to have high prediction capabilities for *IDH* mutation status in high-grade glioma patients”. Despite the great interest in these results, it only includes 25 patients with astrocytoma grade 4. In addition, the molecular basis of these vascular differences between these two high-grade tumors is still unsolved.

## Material and Methods

### Study cohorts

#### Patient cohort with MRI data

The study cohort includes 299 patients with MRI data, 16 of them presented astrocytoma *IDH*-mutant grade 4, and the rest 283 presented glioblastoma *IDH*-wildtype. To collect this cohort, both public (35 patients from TCGA-GBM, 19 patients from Ivy GAP, and 10 from CPTAC-3) and private datasets (108 patients from MTS4UP dataset, 20 from GEINO-mol dataset, and 107 from GLIOCAT dataset) were used. Public datasets are available in The Cancer Imaging Archive (TCIA): https://www.cancerimagingarchive.net/. Private data are available upon reasonable request to the authors. Ethics committee of Universitat Politècnica de València gave ethical approval for this work.

#### Public dataset with RNAseq data

To analyze differences in gene expression between these two types of astrocytoma, two public datasets have been used, including 99 patients from CPTAC3 and 151 patients from TCGA-GBM.

### MRI data

Presurgical MRI studies were collected from all the included datasets, including pre- and post-gadolinium T1-weighted, T2-weighted, FLAIR, and DSC T2*-weighted perfusion sequences obtained by the standard of care protocols using 1.5 or 3.0-T.

#### MRI processing and perfusion markers calculation

To process the MRIs and to calculate the imaging vascular biomarkers, we used the HTS method, included in the ONCOhabitats site (www.oncohabitats.upv.es)^11–13^. The HTS method is illustrated in figure 1 and includes the following stages:

**Figure 1.**
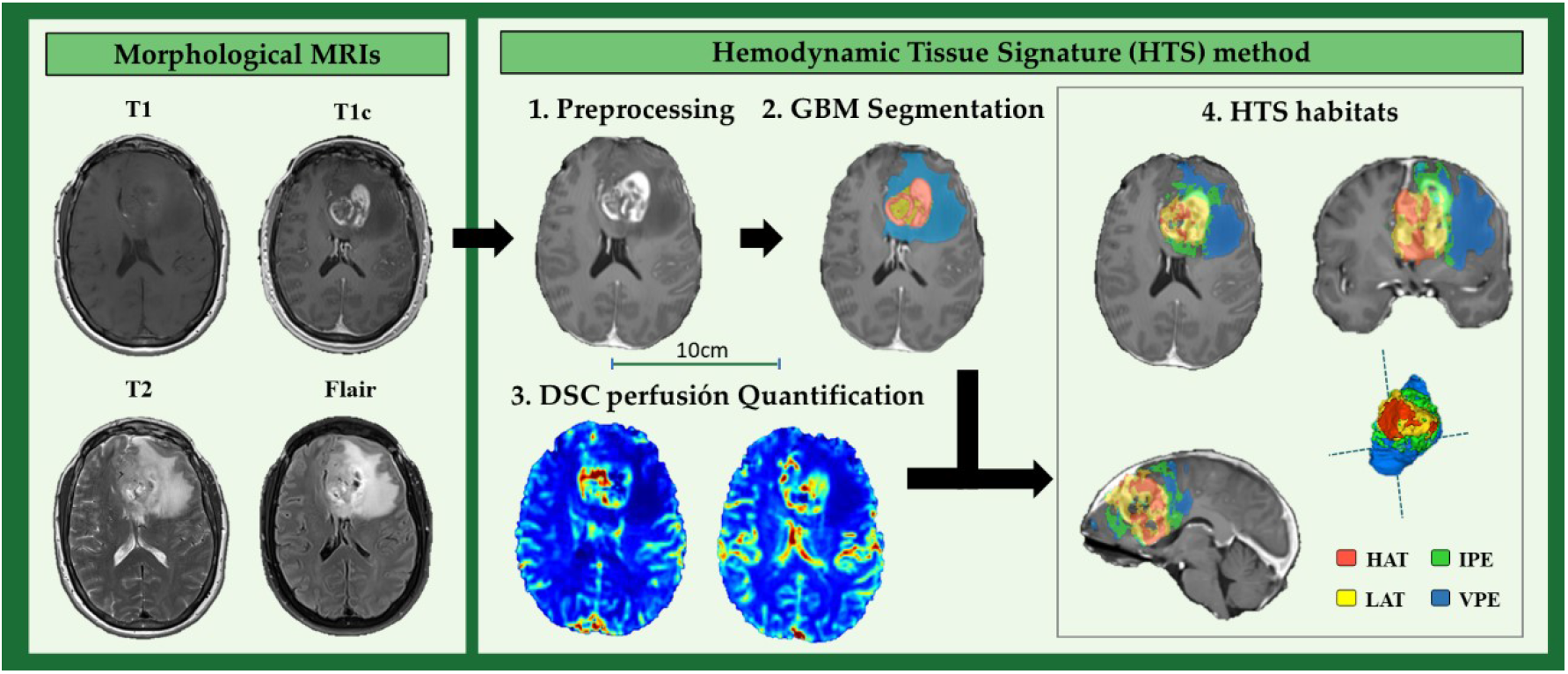
The hemodynamic Tissue Signature (HTS) method, includes the four stages: (1) preprocessing, (2) glioma segmentation in classical tissues: active tumor, edema, and necrosis, (3) DSC perfusion quantification, and (4) HTS habitats: High Angiogenic Tumor (HAT), Low Angiogenic Tumor (LAT), Infiltrated Peripheral Edema (IPE), and Vasogenic Peripheral Edema (VPE).

##### a. Preprocessing

In this stage, common MRI artifacts such as magnetic field inhomogeneities and noise, multimodal registration, brain extraction, or motion correction are corrected.

##### b. Glioma segmentation

A state-of-the-art deep learning 3D convolutional neural network is implemented to segment the enhancing tumor, the edema, and the necrotic tissue. It is based on the directional class adaptive spatially varying finite mixture model (DCA-SVFMM), which is a clustering algorithm that combines Gaussian mixture modeling with continuous Markov random fields to make use of the self-similarity and local redundancy of the images. This methodology uses the unenhanced and GBCA-enhanced T1-weighted sequences, the T2-weighted sequence, and the fluid-attenuated inversion-recovery T2-weighted sequence combined with atlas-based prior knowledge of healthy tissues to delineate the regions.

##### c. DSC perfusion quantification

During this stage, the hemodynamic maps derived from the DSC perfusion sequence are calculated, including Rcbv, rCBF, MTT, and K2 permeability. All perfusion maps are normalized against contralateral unaffected white matter volume to achieve consistency and comparability across different datasets. The normalization is performed automatically by a convolutional neural network, which detects the contralateral unaffected white matter region with 90% accuracy. To ensure a correct perfusion quantification and to avoid under- and over-estimation of perfusion marker, DSC perfusion quantification includes a correction for contrast agent leakage effects. The HTS method implements the Boxerman leakage-correction method^14^ for T1- and T2-leakage effects, as well as gamma-variate fitting to remove the extravasation phase and second pass of the contrast bolus.

##### d. Hemodynamic Tissue Signature (HTS) habitats

In this final stage, an automated unsupervised segmentation algorithm performs the detection of the four vascular habitats within the tumor and edema. Each delineated habitat presents its specific hemodynamic behavior. They are named as: the High Angiogenic Tumor (HAT) habitat, the Low Angiogenic Tumor (LAT) habitat, the Infiltrated Peripheral Edema (IPE) habitat, and the Vasogenic Peripheral Edema (VPE) habitat. HTS habitats are delineated using a DCA-SVFMM structured clustering of rCBV and rCBF maps. The clustering includes two stages: (I) a two-class clustering of the whole enhancing tumor and edema regions and (II) a two-class clustering performed by using only the rCBV and rCBF data within the regions obtained in the first stage. To ensure the reproducibility of the HTS, both stages were initialized with a deterministic seed method.

### RNA data

#### Data Download and Normalization

For both datasets, we retrieved the HT-Seq mRNA read counts from the TCGA database. Samples without *IDH* status information were filtered out to define two experimental groups: *IDH*-mutant and *IDH*-wildtype. Read counts were filtered by expression, removing genes with low counts across all samples. Normalization factors were calculated for each dataset using the Trimmed Mean of M-values (TMM) method^15^.

#### Differential Gene Expression Analysis

We determined the Differential expressed genes (DEGs) in both datasets by fitting a quasi-likelihood negative binomial generalized log-linear model, implemented in the *edgeR* R package^16^. P values were corrected using the Benjamini-Hochberg procedure to control type I errors when conducting multiple comparisons. Genes having an adjusted p-value lower than 0.05 and an absolute value of log Fold-Change greater than 1 were considered as differentially expressed.

### Statistical Analyses

#### Study cohort description

To evaluate the differences in survival between both groups of patients, Kaplan Meier analyses and Log Rank test were performed. The relationships between demographic and clinical variables, and overall survival (OS) were assessed using independent uniparametric Cox regression models.

#### Correlation between MRI-DSC biomarkers calculated at vascular habitats with IDH mutation

We evaluated the significant correlation between each MRI-DSC biomarker with OS to select the optimal biomarkers to find differences among astrocytoma types.

Differences in the imaging vascular biomarkers between *IDH*-wildtype glioblastoma and *IDH*-mutant astrocytoma grade 4 were assessed with Mann Whitney U test, as well boxplots were performed to illustrate differences.

#### Differences in transcriptome between high-grade astrocytoma types

Results from Differential Expression Analysis were functionally annotated using the Biological Processes ontology from the Gene Ontology database^125,126^. First, we selected the GO terms related to vascular processes by matching a list of vascular key terms (table S1 of Supporting Material). Then, we selected all genes mapping with these matched GO terms as our vascular genes set. Most significant vascular and non-vascular DEGs were plotted in heatmaps to illustrate the differences between expression profiles across experimental groups.

Finally, we assessed Overall functional differences between astrocytoma types by performing a Gene Set Enrichment Analysis (GSEA) on Differential Expression Analysis results using the *fgsea* R package^17^. Genes were pre-ranked according to their log fold-change values. Gene sets with an adjusted p-value < 0.05 were considered significant.

## Results

### Detection of relevant prognostic demographic and clinical variables

We analyzed the correlation between OS with the following demographic and clinical characteristics: age, sex, tumor resection type, tumor location (hemisphere), *IDH* mutation, and *MGMT* methylation. Results are included in figure 2A, resulting in the *IDH* mutation as the variable most correlated with longer survival rates (with the lowest hazard ratio), followed by methylated *MGMT* and total tumor resection.

**Figure 2.**
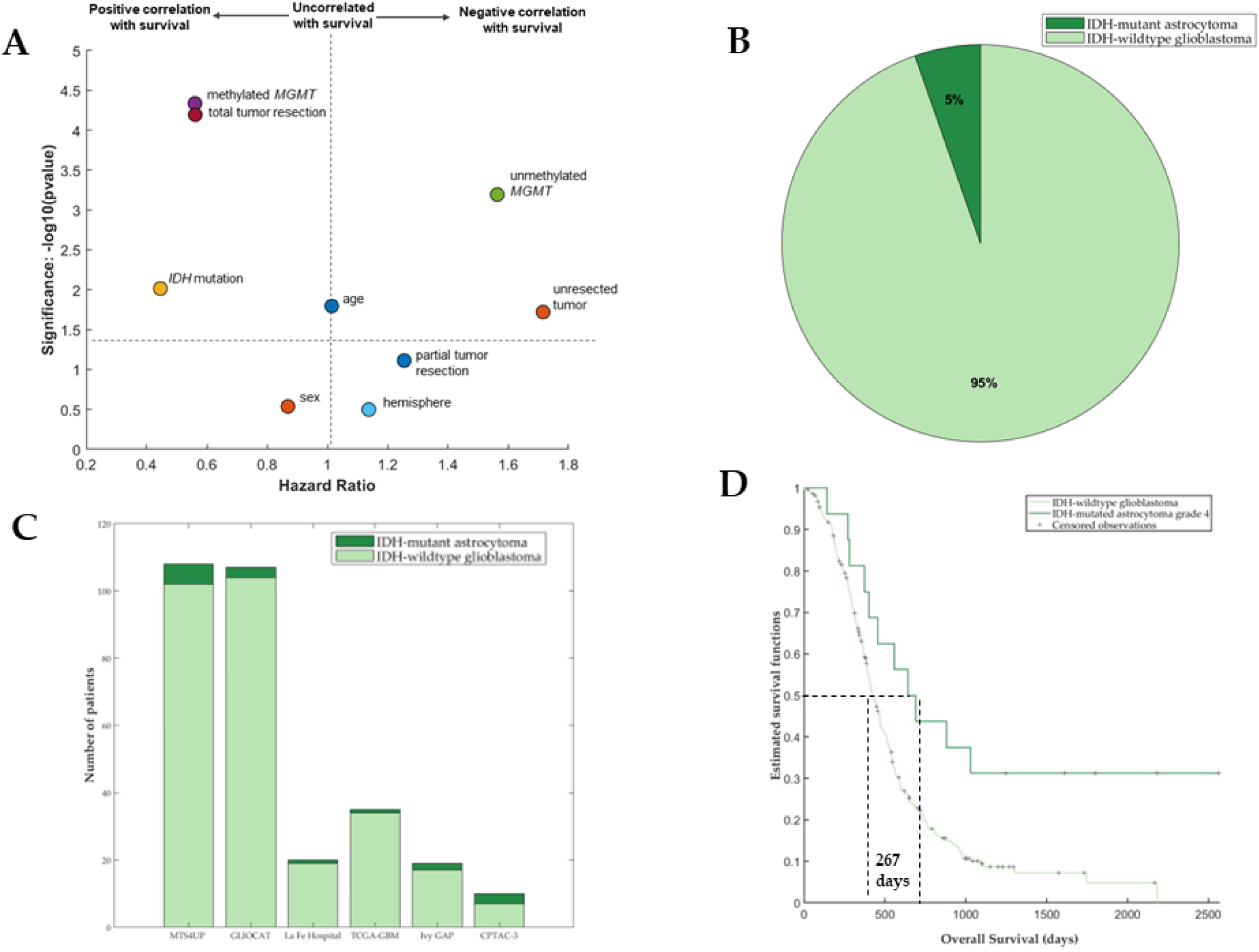
**A)** Scatter plot with the correlation between the main demographic and clinical characteristics and overall survival. **B)** Proportion of patients of the studied cohort with *IDH*-mutant astrocytoma and *IDH*-wildtype glioblastoma. **C)** Number of patients from each dataset with MRI data. **D)** Kaplan Meier curves showing the survival differences between patients with astrocytoma *IDH*-mutant grade 4 (n = 16) and glioblastoma *IDH*-wildtype (n = 287).

### Cohorts’ description

Proportions of each type of astrocytoma in this study dataset are coherent with previously published literature^10,11,118^, representing the population with *IDH*-wildtype glioblastoma a 95% of the entire cohort (figure 2B). In addition, the number of patients from each dataset with *IDH*-mutant astrocytoma and *IDH*-wildtype glioblastoma were compared, showing these proportions differences among datasets (figure 2C).

Differences in OS between these two groups were studied, demonstrating significantly longer survival times for the group of patients with *IDH*-mutant astrocytoma grade 4 (p = 0.001, Log-rank test). Kaplan Meier curves are included in figure 2D, illustrating a median difference in OS of 267 days among the two populations.

Differences in the main demographic, clinical, and molecular features between the two populations were analyzed and the results are included in table 1.

**Table 1.** Demographic, clinical, and molecular features for the cohort with *IDH*-mutant astrocytoma (n = 16) and for the cohort with *IDH*-wildtype glioblastoma (n = 287). P-values resulting from the Mann-Whitney test are also included.

|  | *IDH*-mutant<br>astrocytoma grade 4 | *IDH*-wildtype<br>glioblastoma | p-values<br>(Mann Whitney<br>test) |
| --- | --- | --- | --- |
| #patients (% of entire cohort) | 16 (5.3%) | 287 (94.7%) | - |
| Sex (% Females) | 56.2% | 35.3% | 0.1339 |
| Mean Age at Diagnosis | 49 | 60 | 0.0007* |
| Resection type (#patients) |  |  |  |
| - Total | 5 | 92 | 0.9183 |
| - Partial | 6 | 122 | 0.6609 |
| - Biopsy | 0 | 24 | 0.2265 |
| - Unknown | 5 | 49 | 0.1106 |
| *MGMT* methylation (#patients; %) |  |  |  |
| -Methylated | 6 (37.5%) | 93 | 0.7032 |
| -Unmethylated | 5 (31.2%) | 111 | 0.5262 |
| -Unknown | 5 (31.2%) | 83 | 0.7746 |

Only the median age of diagnosis was significantly different between groups, being lower for the group of patients with astrocytoma *IDH*-mutant grade 4 (49 vs 60 years old).

### Correlation between imaging vascular biomarkers and overall survival

rCBV and rCBF markers at each vascular habitat (HAT, LAT, IPE, and VPE) and for each metric (mean, median, and maximum) were calculated, resulting in 24 markers. We studied the potential prognostic capacity of these markers, analyzing their correlation with *IDH* mutational status.

Figure 3 shows the correlation coefficient and the significance of each MRI-DSC biomarker with overall survival. We can see that the 12 markers related to the HAT and LAT habitats are the most significantly correlated (p<0.05), but only the rCBV markers yield a coefficient higher than 0.2. The selected markers to develop the following analyses were HAT-rCBV_mean_, HAT-rCBV_median_, HAT-rCBV_max_, LAT-rCBV_mean_, LAT-rCBV_median_ and LAT-rCBV_max._

**Figure 3.**
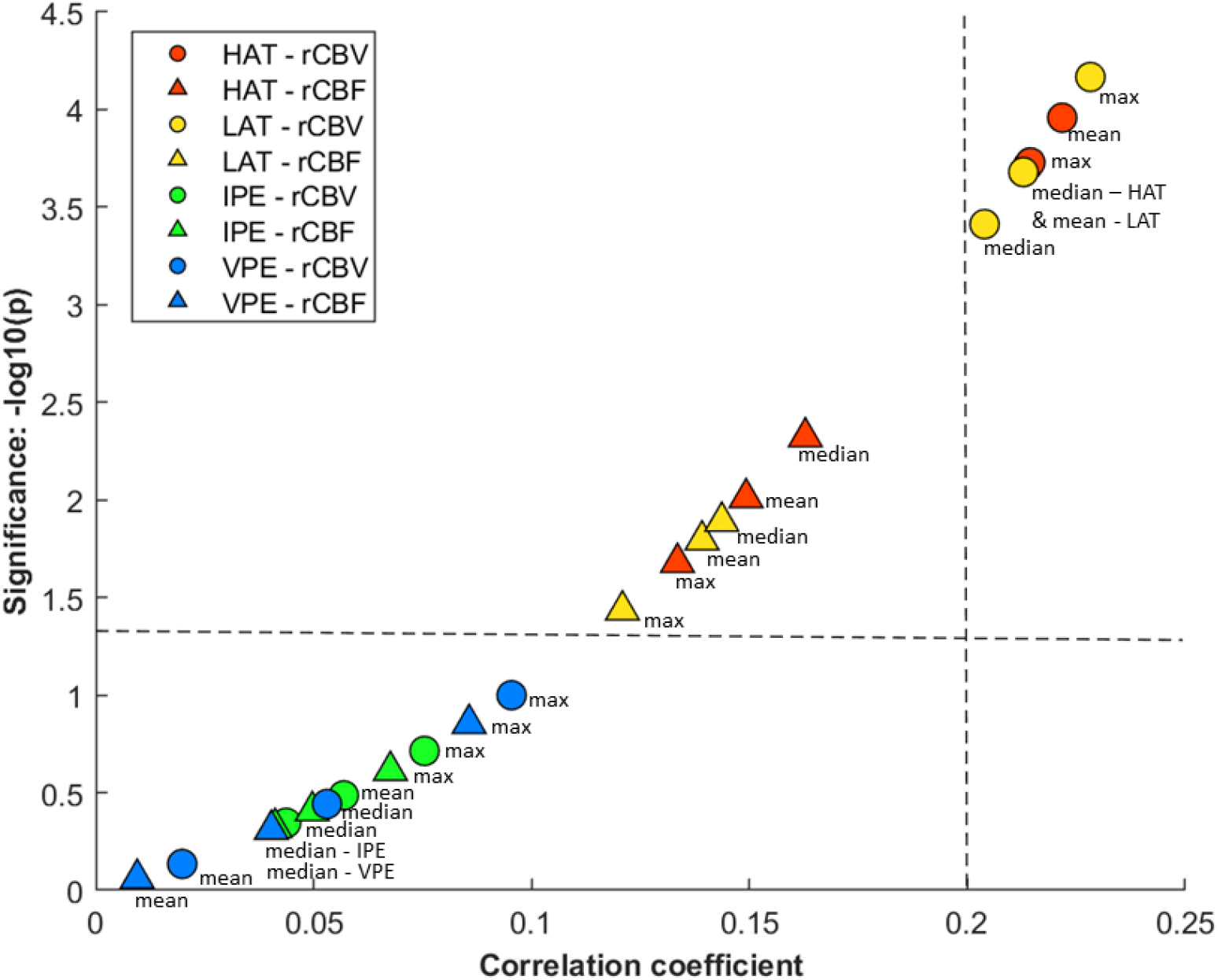
Scatter plot with the correlation results (Spearman coefficients and significance) between MRI-DSC biomarkers and overall survival for the entire cohort (n = 299). Circle markers show rCBV and triangles show the rCBF. Each habitat is represented with a color, HAT in red, LAT in yellow, IPE in green, and VPE in blue, and different metrics are also indicated (mean, median and maximum).

Significant differences (Mann Whitney, p<0.05) in the selected imaging vascular biomarkers between *IDH*-mutant astrocytomas grade 4 and *IDH*-wildtype glioblastomas were found and are illustrated in figure 4. For all the selected MRI-DSC biomarkers, the median value and the minimum and maximum range were higher for the *IDH*-wildtype glioblastoma group, suggesting significantly higher vascularity for these tumors in comparison with *IDH*-mutant astrocytomas grade 4. Table S2 of Supporting Material includes the exact values of each biomarker for the group of *IDH*-wildtype glioblastoma and *IDH*-mutant astrocytoma grade 4.

**Figure 4.**
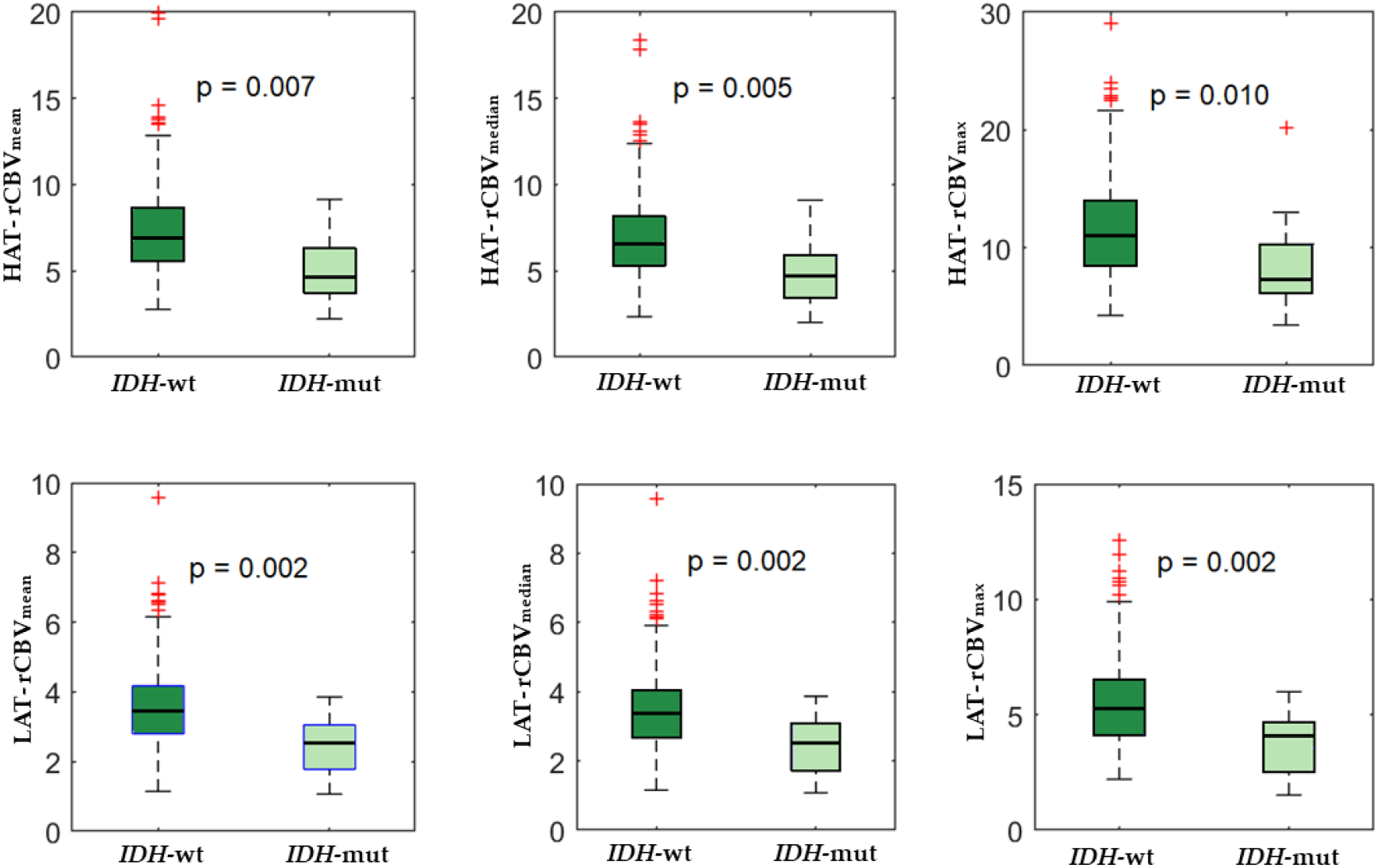
Boxplot showing differences in perfusion between glioblastomas *IDH*-wildtype and astrocytomas *IDH*-mutant grade 4.

### Distinct transcriptome between IDH-wildtype glioblastoma and IDH-mutant astrocytoma grade 4

Differential expression analysis revealed considerable transcriptomic variations between glioblastoma *IDH*-*wildtype* and astrocytoma *IDH*-mutant grade 4. The total number of significant DEGs (adjusted p-value < 0.05 and absolute log Fold-Change > 1) was consistent across datasets (2568 and 2056 DEGs from CPTAC and TCGA, respectively, with an intersection of 879 DEGs).

We detected 1313 (CPTAC) and 1262 (TCGA) overexpressed genes in the *IDH*-wildtype group. A considerable fraction of these DEGs were considered part of the “vascular” functional gene set (143 and 123, respectively, with an intersection of 75 DEGs).

Also, we determined that a total of 1345 (CPTAC) and 794 (TCGA) genes were overexpressed in the *IDH*-mutant group. A total of 30 and 37 DEGs, respectively, were labeled as “vascular” genes, having in common a group of 11 DEGs.

Figure 5 includes the heatmaps showing different gene expressions between glioblastoma *IDH*-wildtype and astrocytoma *IDH*-mutant grade 4 performed with two public datasets. Vascular genes are marked in different colors.

**Figure 5.**
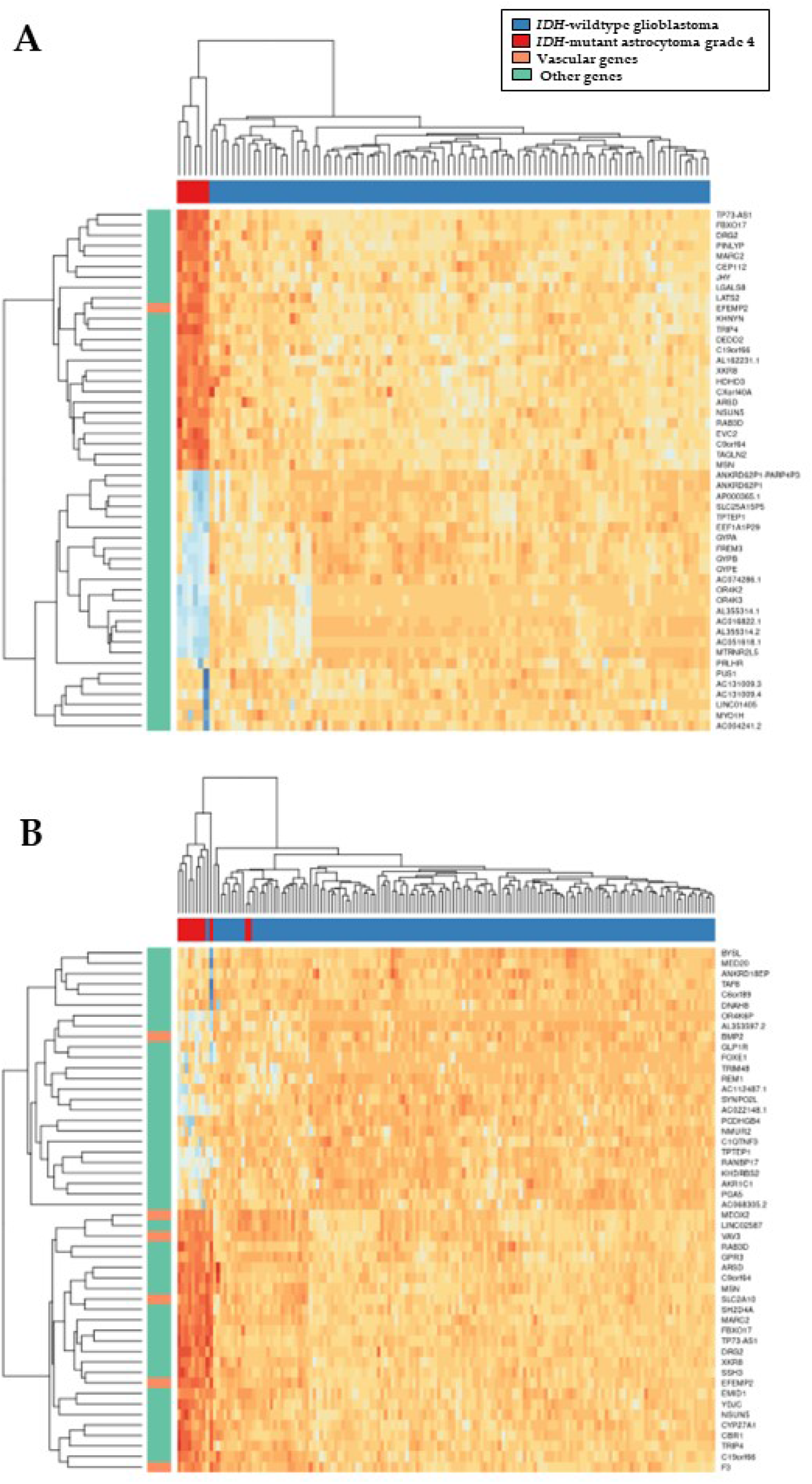
Heatmaps showing different gene expressions between *IDH*-wildtype glioblastoma and *IDH*-mutant astrocytoma grade 4 were performed with two public datasets: **A)** CPTAC3 (n = 93, and 6 respectively) and **B)** TCGA-GBM (n = 140, and 11 respectively)

All significant DEGs, labeled as vascular or nonvascular, are included in the supplementary table S3. This table also contains the adjusted p-values and the log Fold Change values for each gene.

GSEA results showed significant differences between the two groups at the biological processes level. On the one hand, a total of 169 (CPTAC) and 39 (TCGA) GO terms showed significant overrepresentation in IDH-wildtype Glioblastomas. A core subset of 33 GO terms was present in both sets of significant functions.

On the other hand, we only detected 1 (CPTAC), and 3 (TCGA) significantly overrepresented GO terms in *IDH*-mutated Astrocytomas. There was not any coincident function between sets.

## Discussion

With this study we have demonstrated that *IDH*-wildtype glioblastoma and *IDH*-mutant astrocytoma grade 4 present different vascular patterns both in high- and low-angiogenic tumor habitats, defined by the automatic ONCOhabitats method. These differences can be detected using MRI-DSC biomarkers, such as rCBV and rCBF calculated from the presurgical stage, since they are correlated with the presence of the *IDH* mutation. These results are obtained with a multicenter cohort of 299 patients and validate those found in a previous study^10^.

The association between tumor and edema vascularity and OS has been previously analyzed in literature^12,13,18^. There is a consensus that higher values of vascularity, measured by DSC perfusion biomarkers, result in shorter survival times. These important differences in survival may be due in part to different vascular behavior, resulting in quicker progression and greater aggressiveness of *IDH*-wildtype glioblastomas^19–21^. With this work, we have validated this hypothesis, demonstrating significantly higher rCBV values for *IDH*-wildtype glioblastomas. The clinical relevance of this finding is that opens the possibility to use the MRI-DSC biomarkers as complementary tools to support the first diagnosis made by the radiologist from the presurgical stage.

In addition, to acquire a deeper understanding of vascular differences between these two high-grade astrocytomas, we analyzed gene expression patterns of two public datasets that include RNAseq data. We focused on analyzing genes related to vascular processes and structures. We found that 75 genes related to vascularity were up-regulated in both datasets for the group of *IDH*-wildtype glioblastoma, versus only 11 vascular genes up-regulated in the group of *IDH*-mutant astrocytomas grade 4. This finding supports our hypothesis, that stronger vascular processes occur during the establishment and progression of *IDH*-wildtype tumors.

In glioblastomas *IDH*-wildtype, we found overexpression of several genes previously reported as associated with vascular remodeling and arterial abnormalities in high-grade gliomas. Some of these genes are *EGF-containing fibulin-like extracellular matrix protein 2* (*EFEMP2)*^22^, *solute carrier family 2 member 10 (SLC2A10)*^23^, *midkine (MDK)*^24^, and *transmembrane BAX inhibitor motif-containing 1* (*TMBIM1)*^25^. They also have been proven as associated with poor progression, which seems coherent since a higher vascular supply allows quicker tumor progression.

On the other hand, there were 11 vascular genes overexpressed in astrocytomas *IDH*-mutated grade 4 compared with glioblastomas *IDH*-wildtype. We can remark on the relevance of *bone morphogenetic protein 2 (BMP2)*, which has been previously defined as relevant for prognosis estimation^26^, since the higher the expression of *BMP2*, the better the prognosis of patients. In addition, we found an overexpression in *STOX1*, which was previously associated with younger patients and with gliomas of lower grades and less aggressive^27^. The correlation of higher expression of these genes with the presence of *IDH* mutation can be due to the metabolic reprogramming that suffers tumor cells with this alteration.

## Conclusions

In conclusion, we have validated the association between the *IDH* mutation with tumor vascularity, measured by DSC perfusion biomarkers at HAT and LAT habitats. Astrocytomas *IDH*-mutant grade 4 present lower rCBV and rCBF, and longer survival times, which can be partly explained by a slower tumor progression, due to less vascularity.

We have assessed on the potential relevance of specific genes overexpressed in glioblastomas *IDH*-wildtype, such as *EFEMP2, SLC2A10, MDK*, and *TMBIM1*, and in *IDH*-mutant astrocytomas grade 4, such as *BMP2* and *STOX1*. Although it is needed deep analyses, these genes can be proposed as targets for specific novel therapies for each type of high-grade astrocytoma.

This constitutes the first study analyzing vascular differences both at MRI and transcriptomic levels between *IDH*-wildtype glioblastoma and *IDH*-mutant astrocytoma grade 4. We propose DSC biomarkers automatically calculated with the ONCOhabitats method as useful to estimate the *IDH* mutation status in astrocytomas grade 4 from the presurgical stage. Moreover, we suggest the key role of specific genes overexpressed in glioblastomas *IDH*-wildtype as determinant for presenting stronger vascularity; and the clinical relevance of *BMP2* and *STOX1* for astrocytomas *IDH*-mutant grade 4.

## Data Availability

All public data produced are available online at The Cancer Imaging Archive (TCIA): https://www.cancerimagingarchive.net/.
Private data are available upon reasonable request to the authors.

## Funding Information

This study was funded by the Agencia de Investigación de España and framed at the ALBATROSS project: PID2019-104978RB-I00/AEI/10.13039/501100011033

## Supporting Material

**Table S1.** Vascular key terms selected to filter genes of interest.

| Vascular terms |
| --- |
| vascular |
| vasculature |
| blood vessel |
| microvessel |
| angiogenesis |
| hypoxia |
| necrosis |
| capillary |
| glomerulus |
| glomerular |

**Table S2.**
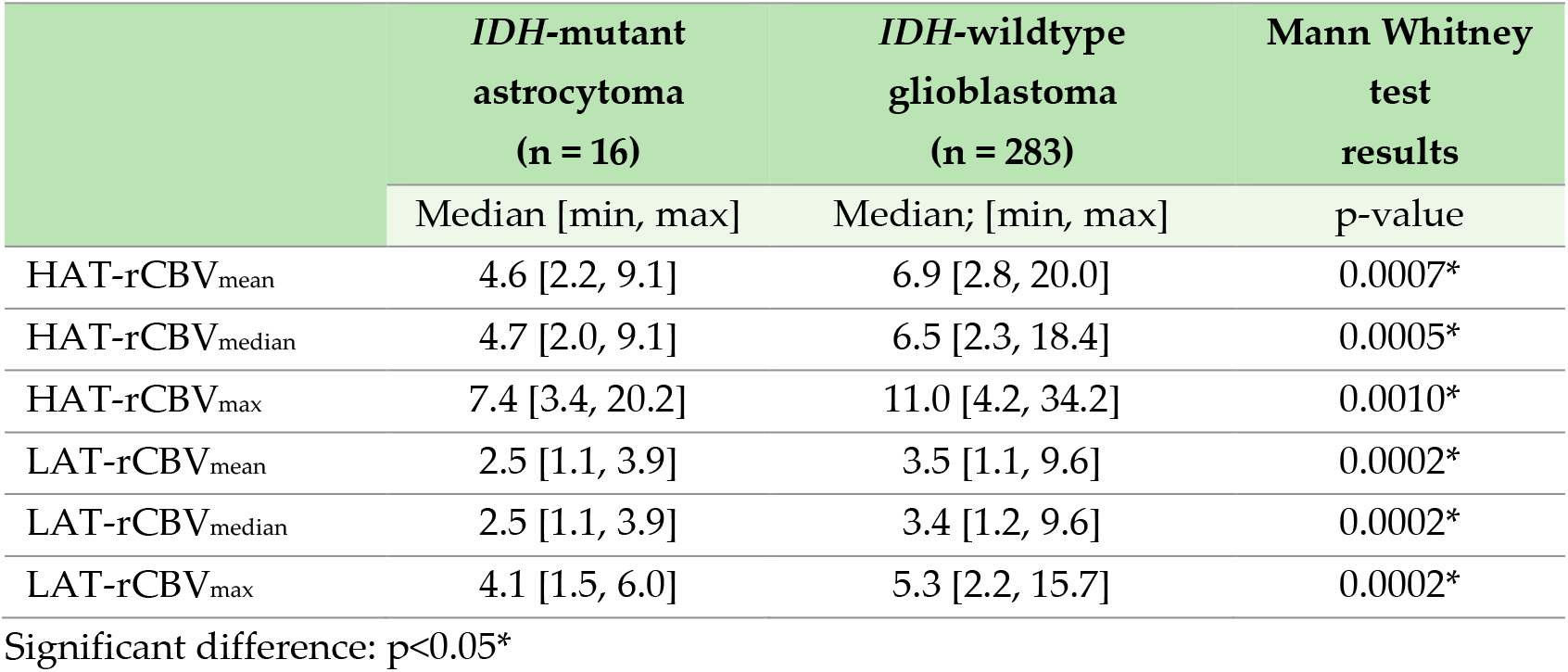
Differences in rCBV between *IDH*-mutant astrocytoma and *IDH*-wildtype glioblastoma, and Mann Whitney results.

|  | *IDH*-mutant<br>astrocytoma<br>(n = 16) | *IDH*-wildtype<br>glioblastoma<br>(n = 283) | Mann Whitney<br>test<br>results |
| --- | --- | --- | --- |
|  | Median [min, max] | Median; [min, max] | p-value |
| HAT-rCBV<sub>mean</sub> | 4.6 [2.2, 9.1] | 6.9 [2.8, 20.0] | 0.0007* |
| HAT-rCBV<sub>median</sub> | 4.7 [2.0, 9.1] | 6.5 [2.3, 18.4] | 0.0005* |
| HAT-rCBV<sub>max</sub> | 7.4 [3.4, 20.2] | 11.0 [4.2, 34.2] | 0.0010* |
| LAT-rCBV<sub>mean</sub> | 2.5 [1.1, 3.9] | 3.5 [1.1, 9.6] | 0.0002* |
| LAT-rCBV<sub>median</sub> | 2.5 [1.1, 3.9] | 3.4 [1.2, 9.6] | 0.0002* |
| LAT-rCBV<sub>max</sub> | 4.1 [1.5, 6.0] | 5.3 [2.2, 15.7] | 0.0002* |

Significant difference: $p < 0.05^*$

## Notes

### Competing Interest Statement

The authors have declared no competing interest.

### Funding Statement

This study was funded by the Agencia de Investigacion de Espana and framed at the ALBATROSS project: PID2019-104978RB-I00/AEI/10.13039/501100011033

### Author Declarations

Ethics committee of Universitat Politecnica de Valencia gave ethical approval for this work.

